# The role of procalcitonin in early differential diagnosis of suspected children with COVID-19

**DOI:** 10.1101/2020.04.07.20057315

**Authors:** Denggao Peng, Jing Zhang, Yingqi Xu, Zhichao Liu, Pengyao Wu

## Abstract

**Background:** We aimed to identify clinical features of coronavirus disease 2019 (COVID-19) in children and evaluate the role of procalcitonin in early differential diagnosis.

**Methods:** A retrospective analysis was performed on all suspected pediatric cases.

**Results:** 39 (50.6%) of 77 suspected cases were comfirmed, 4 (5.2%) of them had viral coinfection. Compared with non-COVID-19 group (n=33), COVID-19 confirmed group (n=39) had fewer fever(OR[95% CI]0.467[0.314-0.694]; P=.000) and symptoms of acute respiratory infection (0.533[0.36–0.788]; P=.001), more asymptomatic (13.568[1.895-96.729]; P=.000), and more family cluster infections (5.077[2.224-11.591]; P=.000), while computed tomography had more positive findings of viral pneumonia (1.822[1.143-2.906]; P=.008). Age (6.9[3.6-10.5] vs 5[2.1-7.6]; P=. 088) and gender were statistically insignificant. Procalcitonin (0.05[0.029-0.076] vs 0.103[0.053-0.21]; P= 000) of COVID-19 alone group (n=35) was significantly reduced. While compared with coinfection group (n=4), procalcitonin (0.05[0.029-0.076] vs 0.144[0.109-2.26]; P=.010) was also reduced. The area under curve of model is 0.834 ([95% CI][0.741-0.926]; P=.000). Procalcitonin as a differential indicator of COVID-19 alone, its area under curve is 0.809 ([0.710-0.909]; P=.000). The optimal cut-off value is 0.1 ng/mL, the sensitivity, specificity, positive and negative predictive value of differentiating are 65.9%, 78.6%, 82.9%, and 59.2%, respectively.

**Conclusions:** Asymptoms or mild symptoms, positive computed tomography findings and family cluster infection are the main clinical features of COVID-19 in children. With good performance, procalcitonin can provide an important basis for differentiating COVID-19 alone and other viral infection or viral coinfection.

## Introduction

Children are naturally susceptible to various respiratory viruses due to their imperfect immune systems. Since the outbreak of coronavirus disease 2019 (COVID-19) began in Wuhan city, Hubei province, China [1,2], more than 2,000 pediatric cases have been reported nationwide in just over two months [3]. Limited by accuracy of nucleic acid detection [4], relative reagent shortage and non-specificity of imaging findings, early differential diagnosis of suspected pediatric patients is difficult to some extent. Obviously, this has a serious impact on timely triage and the following reasonable treatment.

At present, a growing number of studies have focused on diagnosis and treatment of confirmed cases, but few data are available on clinical characteristics and early identification of suspected pediatric patients with COVID-19 as a special population. As a traditional biomarker, procalcitonin (PCT) have shown superior value in differentiating bacterial and viral infections as well as bacterial coinfections. However, the role of PCT in identifying between viruses and viruses or viral coinfection remain unknown. We aimed to identify the clinical features of COVID-19 in children and evaluate the role of PCT in early differential diagnosis, so as to provide a basis for the follow-up timely and reasonable treatment and effective implementation of prevention and control measures.

## Materials and Methods

### Definition and classification

Children are defined as being less than 18 years old. Referring to the guidelines on the diagnosis and treatment of 2019 novel coronavirus infected pneumonia (the sixth edition draft) issued by the National Health Commission of China [5]. Suspected cases are defined as having a clear epidemiological exposure, with or without clinical manifestations, and with or without positive computed tomography (CT) findings. Epidemiological exposure includes close contact with confirmed cases, and or living or traveling in endemic areas (especially Hubei province), and or presence of confirmed case in their residential communities. If nasopharyngeal swab or anal swab specimens tested positive for nucleic acid using real-time reverse transcriptase polymerase chain reaction (RT-PCR) assay, suspected case is identified as a confirmed case. Fever was recognized when body temperature is higher than or equal to 37.3. Symptoms of acute respiratory infection (ARI) includes nasal congestion, runny nose, sneezing, sore throat, cough, expectoration, chest pain, dyspnea, etc. All chest CT images were reviewed by two experienced pediatric radiologists. If unilateral or bilateral lung fields have any of the features as follows:(a) ground glass opacities;(b) consolidations with surrounding halo sign;(c) nodules;(d) residual fiber strips. The result is defined as positive CT findings of viral pneumonia [6]. Family cluster infection was defined as the occurrence of any of the following criteria in 2 or more family members within a period of less than 1 week: (a) fever;(b) symptoms of ARI;(c) positive CT findings of viral pneumonia.

### Data collection

We conducted a retrospective study on all clinical and laboratory data. From Jan 22nd to Mar 1st, all suspected pediatric patients were admitted to the Third People’s Hospital of Shenzhen, and relevant examinations were completed as routine precedures, and clinical and laboratory data of the first day after admission were collected. Based on laboratory pathogen identification results including 2019 novel coronavirus, influenza A, B (INF A, INF B), respiratory syncytial virus (RSV), mycoplasma pneumoniae (MP), and bacteria, all cases were divided into the COVID-19 confirmed group and the non-COVID-19 group. COVID-19 confirmed group was further divided into COVID-19 alone and coinfection group according to whether they were coinfected, and the differences between the groups were compared.

Inclusion criteria: all suspected pediatric cases.

Exclusion criteria of the non-COVID-19 group:(a) pathogen identified as bacteria or MP; (b) coinfection; (c) PCT > 0.5 ng/mL [7].

### Statistical Analysis

All analyses were conducted by using of IBM Statistical Product and Service Solutions software Version 24 (SPSS Inc, Chicago, IL). Continuous variables were summarized as the median with their interquartile ranges (IQRs) or mean with standard deviations (SDs), median [IQR] or [mean ± SD], depending on whether their distributions were normal or not. Comparisons of categorical variables were performed using the Pearson Chi square test. Odds ratios (ORs) and 95% confidence intervals (CIs) were calculated for statistically significant variables. The parametric tests (independent sample Student t-test or One-way analysis of variance) or non-parametric tests (Mann-Whitney U test or Kruskal-Wallis test) were used to analyze variables. The P value was adjusted by the Bonferroni method for comparison between groups. Variables with P <0.2 in the laboratory data analysis were entered into a multivariate binary logistic regression model. Model fitness was assessed with the Hosmer-Lemeshow goodness-of-fit test. Analysis of the area under curve (AUC) of receiver operating characteristic curve (ROC) was constructed to assess the differentiating performance. Sensitivity, specificity, positive predictive value (PPV), negative predictive value (NPV) were also determined. P <.05 was considered as statistically significant in all tests if applied.

## Results

From Jan 22nd to Mar 1st, a total of 77 suspected pediatric patients were admitted, and 39 (50.6%) were confirmed with COVID-19, including 3 (3.9%) cases of RSV coinfection, and 1 (1.3%) case with INF B coinfection.5 (6.5%) of 38 (49.4%) cases of non-COVID-19 were excluded by laboratory results and CT scan, consisting of 3 (3.9%) cases with PCT greater than 0.5 ng/mL, were considered as bacterial infection, and 2 (2.6%) cases were considered as MP coinfection (Supplement Table 1). The including 33 (42.9%) patients consisted of 3 (3.9%) cases of INF A, 2 (2.6%) cases of INF B, 9 (11.7%) cases of RSV and 19 (18.2%) cases of unidentified non-bacterial pathogens.

Compared with non-COVID-19 group (n=33), COVID-19 confirmed group (n=39) had fewer fever(OR[95% CI]0.467[0.314-0.694]; P=.000) and symptoms of ARI (0.533[0.36-0.788]; P=.001), more asymptomatic (13.568[1.895-96.729]; P=000), and more family cluster infections (5.077 [2.224-11.591]; P=.000), while chest CT had more positive findings of viral pneumonia (1.822[1.143-2.906]; P=.008). Age (6.9[3.6-10.5] vs 5[2.1-7.6]; P=.088) and gender (43.6% vs 57.6%; P=344) were statistically insignificant (Table 1). Among all of 4 coinfection cases, 4 had fever and symptoms of ARI, 3 had positive CT findings and 1 was family cluster infection (Supplement Table 1). Compared with non-COVID-19 (n=33) and coinfection (n=4) group, COVID-19 alone (n=35) group had significant statistical differences in PCT (0.05 [0.029-0.076] vs 0.103[0.053-0.21] and 0.144[0.109-2.26]; P=000, P=010), percentage of neutrophil (N%) (40.9[30-48.5] vs 54.8[41.4-67.5]; P=.003), percentage of lymphocyte (L%) (45.3[39.2-56.6] vs 33.2[22.7-42.5]; P=.003), neutrophil count (NC) (2.25[1.43-3.53] vs 3.33[2.8-5.18]; P=.014), lymphocyte count (LC) (2.68[1.99-4.01] vs 2.26 [1.67-2.92]; P=.043), high sensitivity C-reactive protein (hs-CRP) (5.5[1.1-9] vs 9.1 [7.4-9.5]; P=.007). while white blood cell (WBC) count, hemoglobin (Hb) and platelet (PLT) were statistically insignificant (Table 2).

**Table 1:**
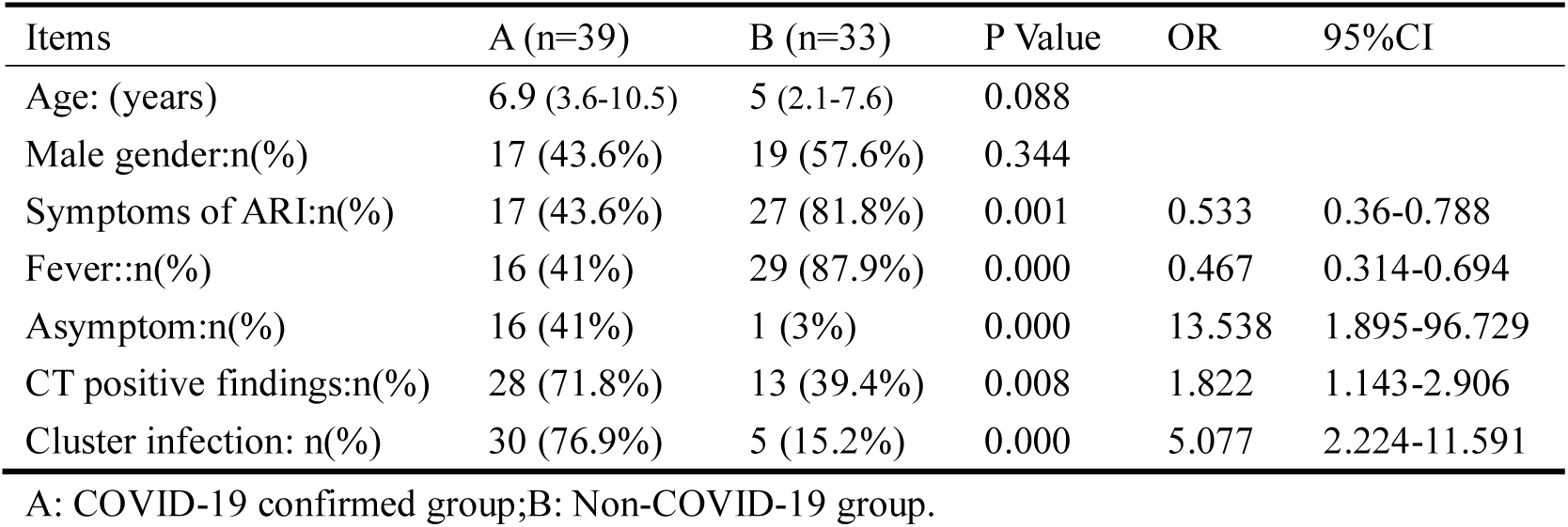
Clinical data analysis between COVID-19 confirmed and non-COVID-19 group

**Table 2:** Comparison of laboratory data between the three groups

| Items | COVID-19 alone<br>(n=35) | Non-COVID-19<br>(n=33) | Coinfection<br>(n=4) | Adjusted P Value |
| --- | --- | --- | --- | --- |
| WBC($\times 10^9/L$) | 5.37 (4.67-8.1) | 6.94 (5.72-8.87) | 8.76 (6.34-13.7) | 0.147 |
| N % | 40.9 (30-48.5) | 54.8 (41.4-67.5) | 34.4 (29.1-45.2) | 0.003,0.003*,1.000** |
| L% | 45.3 (39.2-56.6) | 33.2 (22.7-42.5) | 52.1 (42.9-61.1) | 0.002,0.003*,1.000** |
| NC ($\times 10^9/L$) | 2.25 (1.43-3.53) | 3.33 (2.8-5.18) | 2.78 (2.66-4.6) | 0.018,0.014*,1.000** |
| LC ($\times 10^9/L$) | 2.68 (1.99-4.01) | 2.26 (1.67-2.92) | 5.19 (2.84-7.16) | 0.027,0.043*,0.273** |
| Hb (g/L) | 128 (124-142) | 130 (119-137) | 119 (112-144) | 0.457 |
| PLT ($\times 10^9/L$) | 271.5 (234-328) | 255 (219-314) | 281(202.8-531.8) | 0.204 |
| hs-CRP (mg/L) | 5.5 (1.1-9) | 9.1 (7.4-9.5) | 12.3 (3.2-26.2) | 0.007,0.007*,0.429** |
| PCT (ng/mL) | 0.05(0.029-0.076) | 0.103(0.053-0.21) | 0.144(0.109-2.26) | 0.000,0.000*,0.010** |

\*COVID-19 alone Vs Non-COVID-19; \*\*COVID-19 alone Vs Coinfection

We grouped coinfections and non-COVID-19 infections into one category, and used binary logistic regression analysis to screen independent differential diagnostic indicators for COVID-19 alone infections. WBC, N%, L%, NC, LC, hs-CRP and PCT were entered into a backward stepwise multivariate logistic regression model, and the last step was to obtain two independent indicators of NC (P=0.082) and PCT (P=0.000) (Table 3). Goodness of fit testing (Hosmer-Lemeshow test) was used to assess deviations between observed and expected values. A P value of >.05 implies no significant difference between the observed and expected values. The P value of the goodness-of-fit testing of our model is 0.803, and therefore it is acceptable (Figure 1). Analysis of the AUC of the ROC curve was constructed to assess the differentiating performance. The AUC of overall model is 0.834 ([95% CI][0.741-0.926]; P=.000). PCT as a differential diagnostic indicator of COVID-19 alone, the AUC is 0.809 ([0.710-0.909]; P=. 000) (Figure 1,Table 4). Considering the principle of practicability and good accuracy, the PCT cut-off value is optimized and adjusted to 0.1ng/mL, the sensitivity, specificity, PPV, and NPV of differentiating are 65.9%, 78.6%, 82.9%, and 59.2% (Table 5).

**Figure 1.**
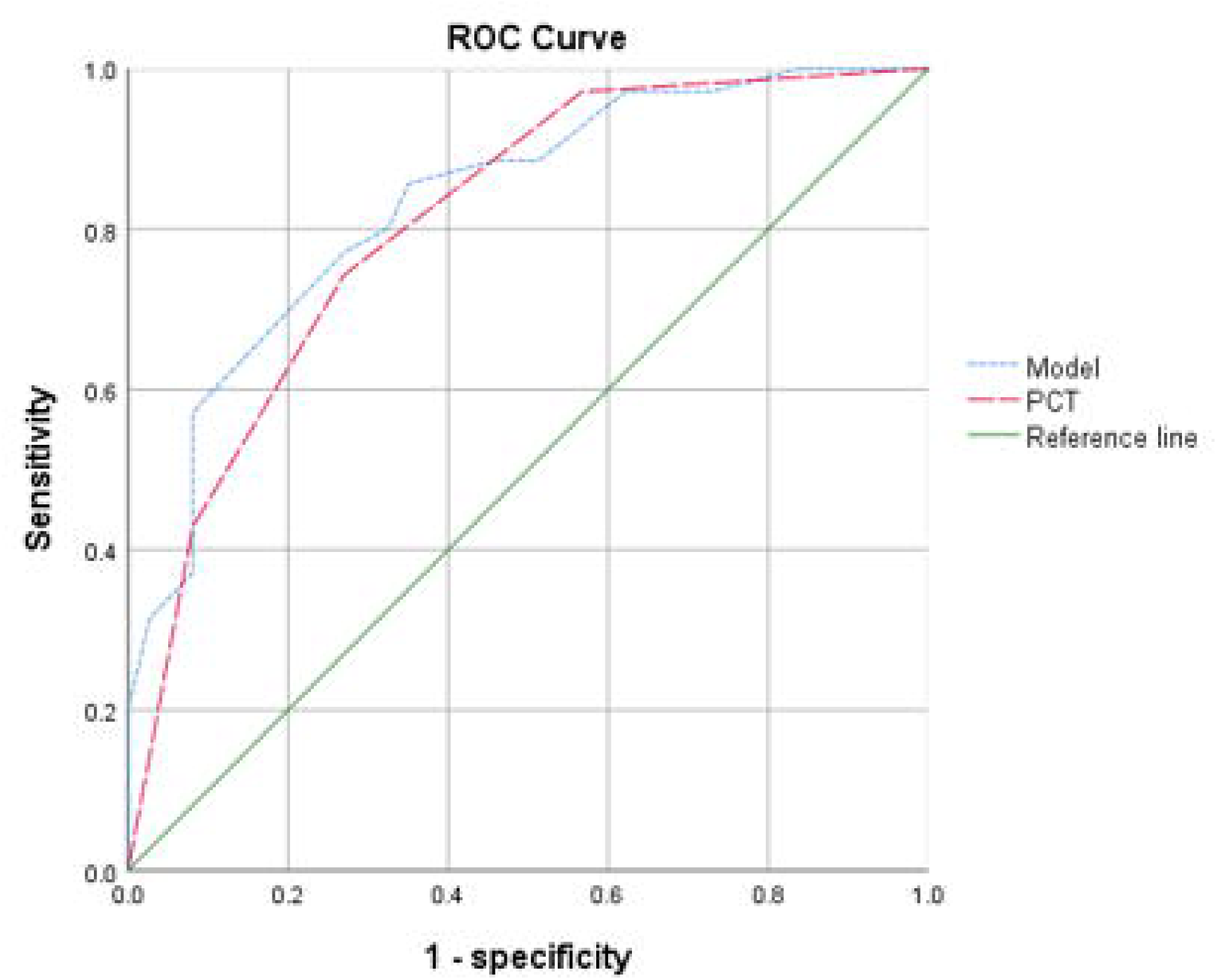
The P value of Hosmer-Lemeshow testing of overall model is 0.803. The AUC for model and PCT were 0.834 and 0.809, respectively.

**Table 3:**
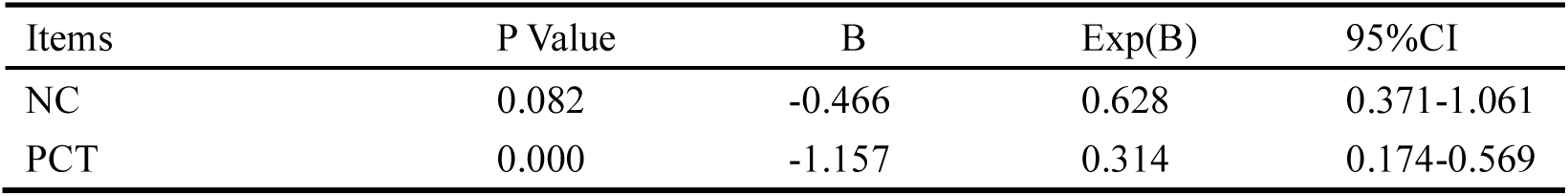
Independent differential diagnostic indicators for COVID-19 alone

**Table 4:**
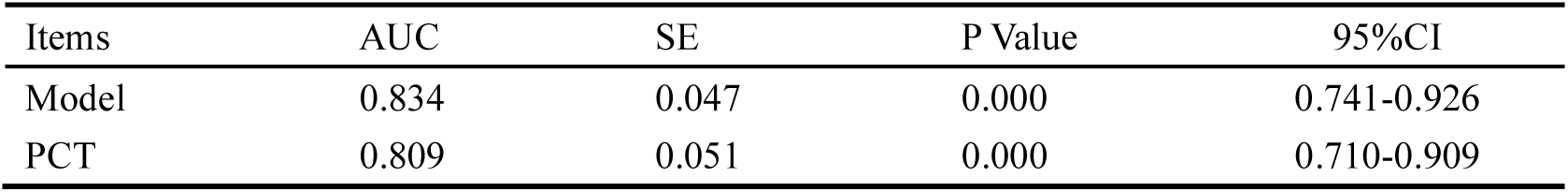
Performance of differential diagnostic indicators for COVID-19 alone

**Table 5:**
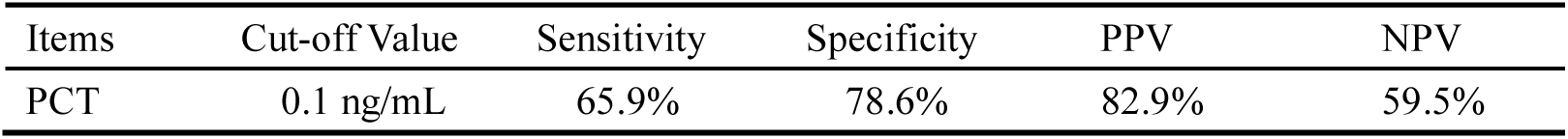
Differentiating characteristics of PCT for COVID-19 alone

| Items | Cut-off Value | Sensitivity | Specificity | PPV | NPV |
| --- | --- | --- | --- | --- | --- |
| PCT | 0.1 ng/mL | 65.9% | 78.6% | 82.9% | 59.5% |

## Discussion

Every winter and spring, a variety of virus infections are prevalent worldwide, such as parainfluenza, RSV, INF A and B, rhinovirus and cytomegalovirus, etc [8,9]. A significant number of community acquired pneumonia (CAP) are caused by viruses, either directly or as part of a coinfection. The clinical picture of these different pneumonias can be very similar, but viral infection is more common in the pediatric populations. Despite great advances in virus detection and pneumonia imaging [4], the identification and differential diagnosis of viral infections has been facing enormous challenges because laboratory detections and radiographic images are not always in agreement with clinical features [10,11]. RT-PCR methods based on spike gene and N gene were widely used for detecting nucleic acid, and were considered a gold standard for COVID-19 confirmed [12-14]. However, this method had its limitations, such as false positive or false negative results, time consuming, false sampling, inconsistence of sample collections and preparations. Most importantly, the inaccuracy of RT-PCR method caused problems in timely triage, isolating the source of infection, the following reasonable treatment, early removal of isolation, and improvement of psychological stress in children. In addition, CT changes in novel coronaviral pneumonia are nonspecific and difficult to differentiate from other viral infections [6,11]. Therefore, it is necessary to start from other clinical features and laboratory data, independent of nucleic acid detection and chest CT, to provide a basis for early differential diagnosis.

As a viral infection, it is obvious that children are especially susceptible to COVID-19. Most of the infected children we observed are asymptomatic or mildly febrile and or symptoms of ARI [3,15]. However, the changes of chest CT are very typical of viral pneumonia [11], which is consistent with the results of other relevant reports. Also, from the COVID-19 cases with coinfection, they all had fever and symptoms of ARI, chest CT findings of viral pneumonia were also typical. Therefore, COVID-19 with other viral coinfections may be more common in children [6]. Pediatric patients should be alert to the possibility of coinfection if severe symptoms appear. Compared with other non-bacterial pathogens, COVID-19 has more family cluster infections, which indicates that the virus is more infectious and has the ability of sustained human-to-human transmission. When developing preventive and control measures for children, disinfecting droplets and household environment and hand hygiene may be the top priorities [16].

For the last two decades, most of studies have been carried out by using, alone or in combination, WBC count, and serum hs-CRP and PCT concentration [17,18]. PCT appears to be most effective in selecting bacterial cases and assessing severity. However, the precise cut-offs between the separation of bacteria from viruses and the separation between mild and severe separations have not been established [19]. In adults, the normal reference value of PCT is less than 0.1 ng/mL. Between 0.1 and 0.25 ng/mL represents a viral infection. Between 0.25 and 0.5 ng/mL, bacterial or bacterial coinfection is less likely to require antibiotic treatment. Data regarding children, despite being limited, are consistent with those collected in adults. Li Z et al found that serum PCT level could provide a useful method of distinguishing bacterial coinfections from an H1N1 influenza infection alone in children during the early disease phase[20]. Chen ZM et al also suggested using a PCT>0.5 ng/mL to identify COVID-19 with bacterial coinfections [7].

Our investigation also used the criterion of PCT>0.5 ng/mL to exclude bacterial infection or coinfection. Three cases in the non-COVID-19 group considered bacterial infections, and one case in the COVID-19 confirmed group had IFN B coinfection, but the possibility of combined bacterial coinfection cannot be ruled out. Interestingly, the PCT of the other three cases with RSV coinfection in the COVID-19 confirmed group was still significantly higher than that of COVID-19 alone. Limited to laboratory testing methods and personnel during the epidemic, except for COVID-19, our hospital can only perform RT-PCR detection of IFN A, B, RSV and cytomegalovirus. Unidentified nonbacterial infections may be other types of viral infections. It also does not rule out that some 2019-nCoV infections may have these other viral coinfection.

There are several limitations in our retrospective cohort study. First, due to the small sample size of the single-center research hospital, in particular, there were only 4 cases in the coinfection group, which may have certain sampling errors. Second, the children may be in different stages of disease when they are admitted to the hospital. Therefore, these results should be carefully interpreted owing to potential selection bias and residual confounding. Larger cohort studies from other cities in China and other countries may also be needed to provide further data support.

## Conclusions

Asymptoms or mild symptoms, positive CT findings and family cluster infection are the main clinical features of COVID-19 in children. With good performance, PCT can provide an important basis for differentiating COVID-19 alone and other viral infection or viral coinfection.

## Data Availability

All data in this paper is true, reliable and available.

## Abbreviations

ARI: acute respiratory infection
AUC: area under curve
CAP: community acquired pneumonia
CI: confidence interval
COVID-19: 2019 novel coronavirus disease
CT: computed tomography
hs-CRP: high sensitivity C-reactive protein
Hb: Hemoglobin
INFA,B: influenza A,B
IQR: interquartile range
LC: lymphocyte count
L%: percentage of lymphocyte
MP: mycoplasma pneumoniae
NPV: negative predictive value
N%: percentage of neutrophil
NC: neutrophil count
OR: odds ratio
PCT: procalcitonin
PPV: positive predictive value
PLT: platelet
ROC: receiver operating characteristic curve
RSV: respiratory syncytial virus
RT-PCR: real-time reverse transcriptase polymerase chain reaction
SD: standard deviation
SE: standard error
WBC: white blood cell.

## Acknowledgements

Thanks for the approval of the Ethics Committee of The Third People’s Hospital of Shenzhen (2020-123).

## Notes

### Competing Interest Statement

The authors have declared no competing interest.

### Funding Statement

Did not receive any specific grant from funding agencies in the public, commercial, or not-for-profit sectors.

